# Development and Validation of a Multivariable Risk Prediction Model for Sudden Cardiac Death after Myocardial Infarction (PROFID Risk Model): Study Rationale, Design and Protocol

**DOI:** 10.1101/2021.07.12.21260002

**Authors:** Glen P. Martin, Gerhard Hindricks, Artur Akbarov, Zoher Kapacee, Le Mai Parkes, Golnoosh Motamedi-Ghahfarokhi, Stephanie Ng, Daniel Sprague, Youssef Taleb, Marcus Ong, Enrico Longato, Christopher A. Miller, Alireza Sepehri Shamloo, Christine Albert, Petra Barthel, Serge Boveda, Frieder Braunschweig, Jens Brock Johansen, Nancy Cook, Christian de Chillou, Petra J.M. Elders, Jonas Faxen, Tim Friede, Laura Fusini, Chris P. Gale, Jiri Jarkovsky, Xavier Jouven, Juhani Junttila, Antti Kiviniemi, Valentina Kutyifa, Daniel Lee, Jill Leigh, Radosław Lenarczyk, Francisco Leyva, Michael Maeng, Andrea Manca, Eloi Marijon, Ursula Marschall, Manickavasagar Vinayagamoorthy, Jens Cosedis Nielsen, Thomas Olsen, Julie Pester, Gianluca Pontone, Georg Schmidt, Peter J. Schwartz, Christian Sticherling, Mahmoud Suleiman, Milos Taborsky, Hanno L. Tan, Jacob Tflt-Hansen, Jan G.P. Tijssen, Gordon Tomaselli, Tom Verstraelen, Kevin Kris Warnakula Olesen, Arthur A.M. Wilde, Rik Willems, Dick L. Willems, Katherine Wu, Markus Zabel, Niels Peek, Nikolaos Dagres

**Affiliations:** Division of Informatics, Imaging and Data Science, Faculty of Biology, Medicine and Health, University of Manchester, Manchester Academic Health Science Centre, Manchester, United Kingdom; Spectra Analytics, London, United Kingdom; Department of Electrophysiology, Heart Center Leipzig at the University of Leipzig, Leipzig, Germany; Department of Information Engineering, University of Padova, Padova, Italy; Division of Cardiovascular Sciences, School of Medical Sciences, Faculty of Biology, Medicine and Health, University of Manchester, Manchester Academic Health Science Centre, Oxford Road, Manchester, United Kingdom; Manchester University NHS Foundation Trust, Manchester Academic Health Science Centre, Southmoor Road, Wythenshawe, Manchester, United Kingdom; Wellcome Centre for Cell-Matrix Research, Division of Cell-Matrix Biology & Regenerative Medicine, School of Biology, Faculty of Biology, Medicine and Health, University of Manchester, Manchester Academic Health Science Centre, Oxford Road, Manchester, United Kingdom; Department of Cardiology, Smidt Heart Institute, Cedars Sinai Medical Center, Los Angeles, CA, USA; Klinikum rechts der Isar, Technische Universität München, Ismaninger Straße 22, D- 81675, Munich, Germany; Cardiology - Heart Rhythm Management Department, Clinique Pasteur, Toulouse, France; Vrije Universiteit Brussel (VUB), Laarbeeklaan 101, 1090 Jette Brussels, Belgium; Paris Cardiovascular Research Center (PARCC), INSERM Unit 970, 56 Rue Leblanc, France; Department of Cardiology, Karolinska University Hospital, Dept of Cardiology Stockholm, Sweden; Department of Cardiology, Odense University Hospital, Department of Cardiology Odense, Syddanmark, Denmark; Division of Preventive Medicine, Brigham and Women’s Hospital, Harvard Medical School, Boston, MA, USA; Département de Cardiologie, CHRU de Nancy, Nancy F-54500, France; Department of General Practice and Elderly Care Medicine, Amsterdam UMC, Vrije Universiteit, Amsterdam Public Health research institute, Amsterdam, the Netherlands; Department of Medical Statistics, University Medical Center Göttingen, Göttingen, Germany; German Center for Cardiovascular Research, partner site Göttingen, Göttingen, Germany; Department of Cardiovascular Imaging, Centro Cardiologico Monzino IRCCS, Milan, Italy; Leeds Institute of Cardiovascular and Metabolic Medicine, University of Leeds, Leeds, United Kingdom; Department of Cardiology, Leeds Teaching Hospitals NHS Trust, Leeds, United Kingdom; Institute of Biostatistics and Analyses, Faculty of Medicine, Masaryk University, Czech Republic; Research Unit of Internal Medicine, Medical Research Center Oulu, University of Oulu and Oulu University Hospital, Oulu, Finland; Biocenter Oulu, University of Oulu, Oulu, Finland; University of Rochester Medical Center, Clinical Cardiovascular Research Center, Rochester, New York, USA; Feinberg Cardiovascular and Renal Research Institute, Northwestern University Feinberg School of Medicine, Chicago, IL, USA; Boston Scientific Corporation, St. Paul, Minnesota, USA; Department of Cardiology, Congenital Heart Defects and Electrotherapy, Medical University of Silesia, Silesian Center of Heart Disease, Zabrze, Poland; Aston Medical School, Aston University, Aston Triangle, Birmingham, United Kingdom; Department of Clinical Medicine and Department of Cardiology, Aarhus University, Aarhus, Denmark; Centre for Health Economics, University of York, York, United Kingdom; European Georges Pompidou Hospital and University of Paris, Paris, France; Department of Medicine and Health Services Research, BARMER Health Insurance, Lichtscheider Strasse 89, 42285 Wuppertal, Germany; Department of Cardiology, Odense University Hospital, Odense, Denmark; Istituto Auxologico Italiano, IRCCS, Center for Cardiac Arrhythmias of Genetic Origin, Milan, Italy; University Hospital Basel, University Basel; Department of Cardiology, Rambam Health Care Campus, Haifa, Israel; Department of Internal Medicine I – Cardiology, Olomouc University Hospital, Olomouc, Czech Republic; Dept of Clinical and Experimental Cardiology, Amsterdam University Medical Center location AMC, Amsterdam, the Netherlands; Netherlands Heart Institute, Utrecht, the Netherlands; The Department of Cardiology, The Heart Centre, Copenhagen University Hospital, Rigshospitalet, Copenhagen, Denmark; Department of Cardiology, Academic Medical Center, University of Amsterdam, Amsterdam, the Netherlands; The Albert Einstein College of Medicine, Bronx, New York, United States; University Hospitals (UZ) Leuven, Leuven; Department of Ethics, Law and Humanities, Amsterdam University Medical Center, University of Amsterdam, Amsterdam, the Netherlands; Division of Cardiology, Department of Medicine, Johns Hopkins University School of Medicine, Baltimore, Maryland, USA; Department of Cardiology and Pneumology, Heart Center, University Medical Center Goettingen, Robert-Koch-Strasse 40, 37075 Goettingen, Germany

**Author notes:** **Corresponding Author** Dr Glen Philip Martin, Senior Lecturer in Health Data Science, Vaughan House, University of Manchester, Manchester, M13 9GB, United Kingdom. G.P.M. and G.H. contributed equally and are considered joint first authors. N.P. and N.D. contributed equally and are considered joint senior authors. **Contributorship statement** Gerhard Hindricks and Nikolaos Dagres conceived the overall idea of the PROFID project and are the study principal investigators. Glen P. Martin, Zoher Kapacee and Niels Peek drafted the initial version of this manuscript, with scientific input on the study design and methodology from Artur Akbarov, Le Mai Parkes, Golnoosh Motmedi-Ghahfarokhi, Stephanie Ng, Daniel Sprague, Youssef Taleb, Marcus Ong, Enrico Longato and Chris Miller. All authors critically reviewed the manuscript for scientific content and have approved the final version. **Competing Interests** The authors have no conflicts of interest to declare. **Funding** The work described is part of the PROFID project. This project has received funding from the European Union’s Horizon 2020 research and innovation programme under grant agreement No 847999.

**Keywords:** Clinical prediction model, model development and validation, sudden cardiac death, myocardial infarction, protocol, defibrillator implantation

## Abstract

**Introduction:** Sudden cardiac death (SCD) is the leading cause of death in patients with myocardial infarction (MI) and can be prevented by the implantable cardioverter defibrillator (ICD). Currently, risk stratification for SCD and decision on ICD implantation are based solely on impaired left ventricular ejection fraction (LVEF). However, this strategy leads to over- and under-treatment of patients because LVEF alone is insufficient for accurate assessment of prognosis. Thus, there is a need for better risk stratification. This is the study protocol for developing and validating a prediction model for risk of SCD in patients with prior MI.

**Methods and Analysis:** The EU funded PROFID project will analyse 23 datasets from Europe, Israel and the US (∼225,000 observations). The datasets include patients with prior MI or ischemic cardiomyopathy with reduced LVEF<50%, with and without a primary prevention ICD. Our primary outcome is SCD in patients without an ICD, or appropriate ICD therapy in patients carrying an ICD as a SCD surrogate. For analysis, we will stack 18 of the datasets into a single database (datastack), with the remaining analysed remotely for data governance reasons (remote data). We will apply 5 analytical approaches to develop the risk prediction model in the datastack and the remote datasets, all under a competing risk framework: 1) Weibull model, 2) flexible parametric survival model, 3) random forest, 4) likelihood boosting machine, and 5) neural network. These dataset-specific models will be combined into a single model (one per analysis method) using model aggregation methods, which will be externally validated using systematic leave-one-dataset-out cross-validation. Predictive performance will be pooled using random effects meta-analysis to select the model with best performance.

**Ethics and dissemination:** Local ethical approval was obtained. The final model will be disseminated through scientific publications and a web-calculator. Statistical code will be published through open-source repositories.

## Introduction

Sudden cardiac death (SCD) is the leading cause of death,[1] accounting for approximately 20% of all deaths in Europe [2,3], and the estimated yearly incidence of SCD in European countries is around 1 per 1,000 inhabitants,[4] afflicting approximately 350,000-700,000 Europeans annually.[3–6] The majority of SCD cases occur in people with coronary artery disease and are mostly caused by ventricular tachyarrhythmias following myocardial infarction (MI). After MI, a reduced left ventricular ejection fraction (LVEF) is associated with increased risk for all-cause death, cardiac death and SCD.[8] Randomised clinical trials (RCTs) have demonstrated that in patients with severely impaired LVEF, the risk of SCD and all-cause death may be significantly reduced through prophylactic implantation of an implantable cardioverter defibrillator (ICD) [9–11]. Based on these data, international guidelines recommend ICD implantation in post-MI patients with severely reduced LVEF ≤35% for primary prevention of SCD.[1]

However, since completion of these landmark trials in the late 1990s and early 2000’s, major advances in pharmacological and non-pharmacological treatment have led to substantial decline in the risk of SCD after MI.[1,3] Concordantly, only a minority of these patients will ever need the implanted ICD and experience appropriate ICD therapies. In fact, the numerical majority of SCD cases occur in patients with LVEF>35% and who are not considered for a primary prevention ICD implantation according to current guidelines [3,12]. Thus, there is consensus that the current practice of using LVEF as the sole risk stratification factor for the risk of SCD after MI and the decision on prophylactic ICD implantation has significant limitations. [13]

Other clinical characteristics, laboratory and imaging biomarkers, genetic markers and risk factors have been reported to be associated with increased risk of SCD.[3,13,14] However, in isolation, none of these prognostic factors have sufficient predictive accuracy for clinical use. Consequently, there is an imperative and unmet need for the development of a clinical prediction model (CPM) to use a combination of such predictor variables to estimate the risk of SCD after MI. Existing CPMs [15–19] have not been implemented into recommended practice, largely because improvements in their predictive ability (and clinical utility [20]) is required.

The development of a CPM for risk of SCD in patients with previous MI is a primary objective of the PROFID project – a large Horizon 2020 funded pan-European consortium [21] (grant no. 847999). The aim of this CPM is to aid the risk stratification of patients with MI and facilitate decision-making for primary prevention ICD implantation. Once the PROFID CPM has been developed, the PROFID project will compare its predictive performance with the existing CPMs (including using LVEF as the sole risk stratification variable, reflecting current clinical practice). The project will then compare personalised decision-making for prophylactic ICD implantation with application of the CPM against current clinical practice in patients with LVEF≤35% and LVEF>35% in two multinational randomised clinical trials, (PROFID-Reduced and PROFID-Preserved, NCT04540354 and NCT04540289).

The aim of this paper is to describe the study protocol for the development and validation of the PROFID CPM, including the key analytical steps and decisions that have been considered.

## Methods and Analysis

We structure this protocol in line with the transparent reporting of a multivariable prediction model for individual prognosis or diagnosis (TRIPOD) statement and checklist.[22] Given that this is a protocol, we focus on the methods section of the TRIPOD statement. Subsequent publications reporting the results will adhere to the full TRIPOD checklist.

### Aim

The aim of the study, on which this protocol is based, is to develop and validate a multivariable CPM to estimate the risk of SCD after MI, using data from Europe, Israel and the US.

### Study Design and Data Sources

This will be a retrospective analysis of data arising from observational cohort studies, routine healthcare records and randomised controlled trials. **Table 1** shows the full list of datasets used for the development of the CPM, comprising 23 datasets from 12 countries. All datasets describe individuals who have had an MI or have coronary artery disease with ischemic cardiomyopathy and reduced LVEF <50%, and all contain information on SCD (or surrogates thereof). We distinguish between four types of datasets: (i) acute MI cohort datasets, where each patient was entered into the dataset at time of their acute MI event; (ii) prior MI or ischemic cardiomyopathy cohort datasets, where patient had a previous MI or ischemic cardiomyopathy and reduced left ventricular ejection fraction (<50% as defined by current heart failure guidelines of the European Society of Cardiology [23]) and were entered into the dataset at some time after their MI event; (iii) ICD cohort datasets, where each patient was entered into the dataset at the time of receiving their prophylactic ICD implant for primary prevention of SCD after an MI; and (iv) datasets from randomised controlled trials in which participants with prior MI or ischemic cardiomyopathy and severely reduced LVEF, received ICD implants or medication therapy (e.g. MADIT II,[9,10], SCD-HeFT [11]). All datasets contain clinical information, medications and other selected variables such as ECG parameters and biomarkers, which were recorded for each patient at time of entry into the dataset. Some datasets include additional variables regarding cardiac magnetic resonance (CMR) imaging, which have been shown to be highly prognostic of SCD [24] (see **Table 1**). All data include information about outcomes during follow-up for each patient: until time of SCD, death from other causes, or until time of first appropriate ICD shocks/anti-tachycardia pacing, hereafter referred to as appropriate therapy.

**Table 1:**
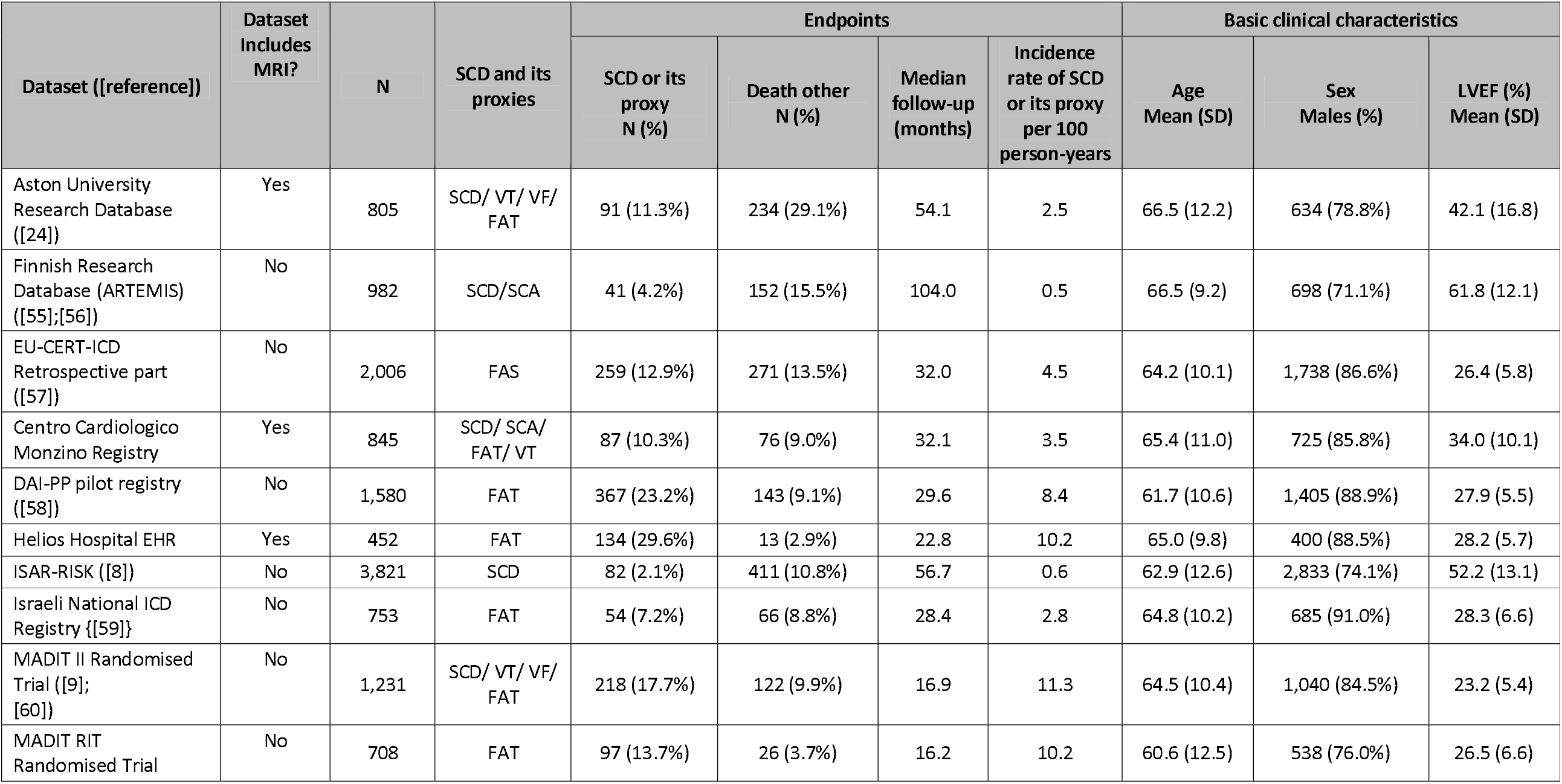
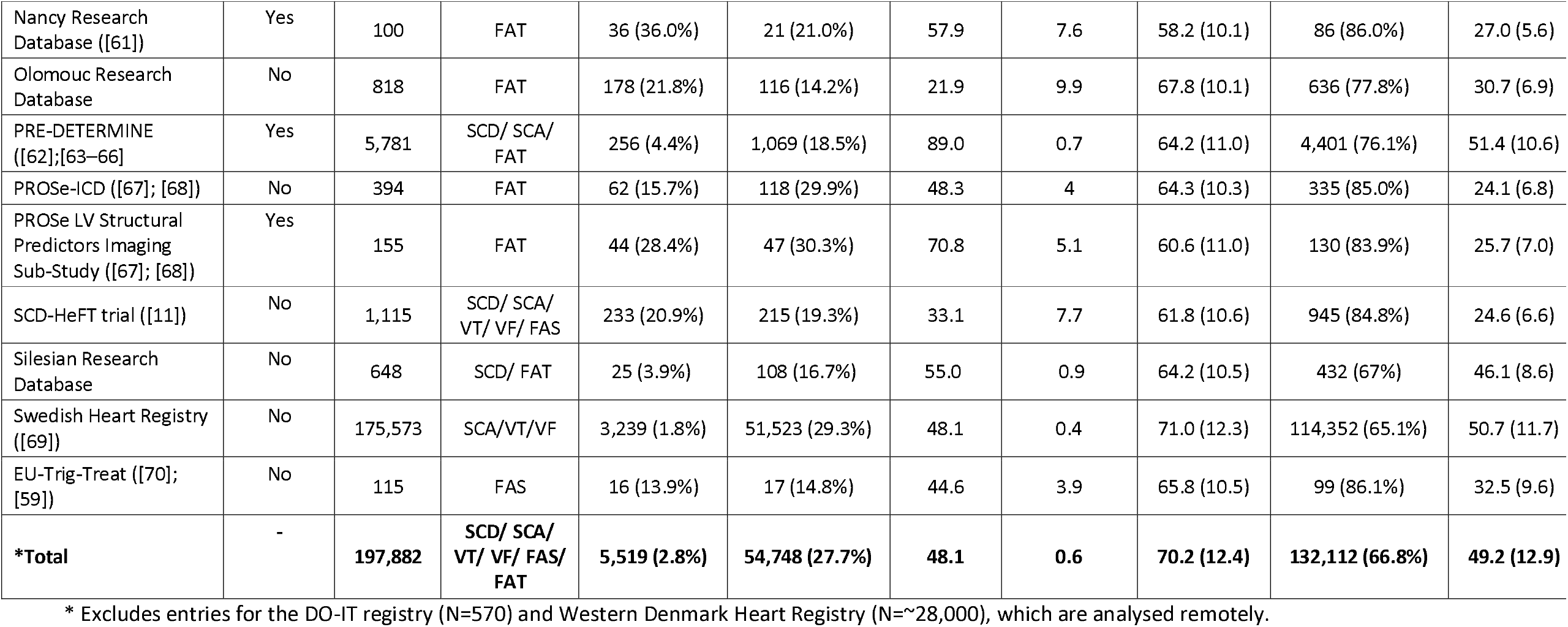
Overview of the datasets included in our study. The population column shows whether the data are of type “acute MI cohort”, “prior MI cohort”, “ICD cohort”, or “randomised controlled trials”, as defined in the methods section.

Prior to analysis, all datasets will undergo data cleaning in partnership and agreement with the respective data providers. Given that each of the datasets contain different variables, we have developed a common data model to ensure that there is a consistent set of variables across all data sources (**Supplementary Table 1**). In particular, the common data model dictates the units of measurement where applicable, categories for nominal and ordinal variables, and definitions of each variable. The variables listed in the common data model define the list of candidate predictor variables we will consider for inclusion in the model (i.e., for variable selection – see supplementary methods). We will also include calendar year in the models as a continuous variable to account for varying risk through time. At the time of prediction, this will be always set to the latest calendar year in the development data.

### Participant Entry Criteria

Our analysis will include all patients who are at least 18 years old and have had either: (a) a previous MI defined as ST-segment-elevation myocardial infarction or non-ST-segment-elevation myocardial infarction, or (b) coronary artery disease and ischemic cardiomyopathy with reduced left ventricular ejection fraction <50% (as defined by current heart failure guidelines of the European Society of Cardiology [23]). From these patients, we exclude: (i) patients who received ICD implantation for secondary prevention of SCD at baseline, (ii) patients with a cardiac resynchronization therapy (CRT) device at baseline, (iii) patients with non-ischemic cardiomyopathy such as dilated cardiomyopathy, hypertrophic cardiomyopathy, or restrictive cardiomyopathy, (iv) patients with a primary electrical arrhythmic disease such as long QT syndrome, Brugada syndrome or catecholaminergic polymorphic VT, (v) patients with congenital heart disease, and (vi) patients who died (or experience the outcome) within the first 40 days after the index MI. A schematic of the inclusion and exclusion criteria is given in **Figure 1**.

**Figure 1:**
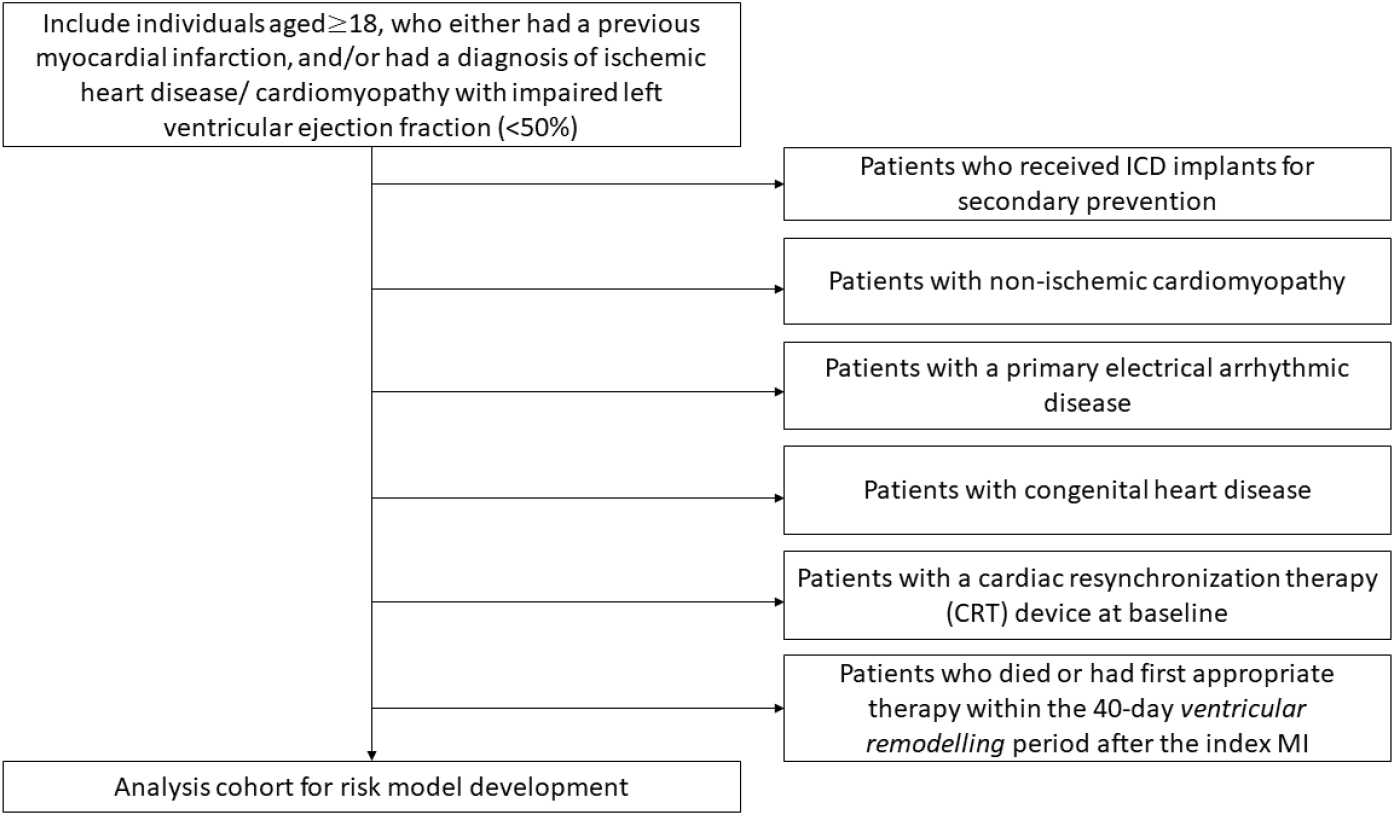
Schematic of the inclusion and exclusion criteria, which are applied to each dataset within the PROFID project.

Patients with a CRT device at baseline are excluded because CRT reduces the risk for SCD [25]. The reason for exclusion criterion (vi) is that, after MI, patients undergo a ventricular remodelling period, during which important parameters for the subsequent risk such as LVEF may significantly change. Therefore, decisions on prophylactic ICD implantation are made after ventricular remodelling. Since the intention is to use the PROFID CPM after ventricular remodelling, we designed the study to match this prediction time point (i.e., predictions are possible any time after the 40-day remodelling period).

### Outcome definitions and analytic approach

Our primary outcome is time-to-sudden cardiac death or to life-threatening ventricular arrhythmias (ventricular tachycardia, or ventricular fibrillation) during follow-up. Time-zero for calculating the time-to-event outcomes will be defined as follows:

1. In the acute MI cohort datasets, time-zero for each patient is 40 days after the index MI event. If an individual has more than one MI event within the dataset, then their index MI will be chosen randomly across all their MI events prior to their first competing risk outcome. The decision to take a random MI event ensures maximum consistency across datasets and avoids introducing biases if one were to select the first or last MI.
2. In the prior MI or ischaemic cardiomyopathy cohort datasets, time-zero for each patient is their time of entry into the dataset/study, provided it is more than 40 days after the initial MI event.
3. In ICD datasets, time-zero for each patient is the time of ICD implantation for primary prevention of SCD, provided it is more than 40 days after the initial MI event.

The primary outcome for each patient is then defined as the time between time-zero and SCD or life-threatening ventricular arrhythmia event, which is defined across our datasets as follows, depending on whether a patient has an ICD implant or not:

- In patients without an ICD, the primary outcome is occurrence of SCD based on cause-of-death adjudication, or implantation of ICD for secondary prevention of SCD (i.e., due to the occurrence of ventricular tachycardia or ventricular fibrillation). The definitions for which cause-of-deaths were listed as SCD varies across datasets. Moreover, the Swedish Heart Registry does not contain information on SCD directly, rather whether the patient had a successful resuscitation outcome for sudden cardiac arrest, which will be used as a surrogate for SCD (i.e., the first resuscitation event will be used as the endpoint for that individual). We therefore expect to see heterogeneity in the incidence of SCD across the included datasets, which will be accounted for directly through the modelling (see below).
- In patients with an ICD, the primary outcome will be defined as appropriate therapy delivered by an ICD. This surrogate for SCD is required since patients with an ICD cannot experience SCD that is preventable by the device. First appropriate therapy mainly includes first appropriate shock and/or the first appropriate anti-tachycardia pacing (ATP); however, some datasets do not contain information on ATP, where first appropriate therapy will include appropriate shock only. We will account for such differences within the models (see below). A limitation of using appropriate therapy is that the programming of each ICD device differs across datasets and patients. However, appropriate ICD therapies remain the best available surrogate for SCD in patients with an ICD, particularly considering the objective of applying the CPM for decision-making on need for ICD implantation.

For any patient who receives an ICD for primary prevention of SCD during their follow-up (i.e., without prior occurrence of ventricular tachycardia or ventricular fibrillation), the data on appropriate therapies after the time of primary prevention ICD implant will be used to define their time-to-event outcome. If a patient receives an ICD during follow-up and we do not have data on subsequent appropriate therapies, then we will censor this patient at the time of ICD implantation, unless this ICD was implanted for secondary prevention (ventricular tachycardia or ventricular fibrillation) in which case it will be taken as our primary outcome; such censoring will be only required in the Swedish Heart Registry.

In all time-to-event analyses, we will account for competing risks of death from other causes using the Fine and Gray competing risks modelling framework.[26] Specifically, in patients without an ICD, we distinguish (i) SCD (resuscitated or non-resuscitated, including sustained VT/VF), and (ii) death from any other cause, where (ii) is a competing risk for (i). In patients with an ICD, we distinguish (i) appropriate therapy (shock – with or without ATP), and (ii) death from any other cause including SCD that was not prevented by an ICD.

Heart transplantation or implantation of left ventricular mechanical assist device during follow-up will be considered a censoring event. All other censoring will be administrative censoring.

The differences in the outcome definition across patients with and without ICDs causes heterogeneity and clustering in our analysis, which we will account for during the modelling by including the following patient-level binary indicators as covariates: (i) entry of study with an ICD vs. entry of study without an ICD, (ii) patient from the Swedish Heart Registry (resuscitation outcomes only) vs. patient not from the Swedish Heart Registry, and (iii) appropriate therapy defined as shock only vs. shock or anti-tachycardia pacing. We will also include interactions in the model between LVEF and the timing of such measurement (before or after 40-day ventricular remodelling period).

### Sample Size calculation

Sample size criteria for the development of CPMs for continuous, binary and time-to-event outcomes have recently been proposed.[27–29]. Since sample size criteria for competing risk models (and non-regression-based models) have yet to be developed, we base our sample size calculation on the criteria developed for time-to-event prediction models.[28] We assumed the following when making our sample size calculations for developing a time-to-event CPM (some values, such as event proportions, are based on the datasets available to us): (i) 2.2% experience SCD during follow-up, (ii) the mean follow-up time is 4.5 years, (iii) the model would explain 15% of maximum R^2^, (iv) we target a maximum degree of shrinkage/overfitting of 0.9, and (v) we have approximately 100 candidate predictors in our common data model. This resulted in a minimum required sample size of 24219, which was driven by criteria 1 of Riley et al.[28] Combined, the PROFID datasets include approximately 225,000 patients, which far exceeds the minimum requirements.

### Missing Data

Many of the datasets in this study will contain variables with missing values. During the exploratory and descriptive analyses, we will investigate missing data patterns using graphical plots and tabulations. During CPM development, validation, and deployment, we will use fuzzy K-means to impute missing data,[30] which has been shown to be a robust method in prediction context.[31] For this study, fuzzy K-means is especially attractive over alternative imputation methods because it can be easily implemented during model deployment since it only requires cluster centroids to be retained (which poses no risk in terms of information governance). While multiple imputation would be an alternative imputation strategy for developing the CPM, it poses issues when deploying the model in practice, because to use the CPM with missing data would require one to store a copy of the development data, and as such will not be considered here. [32] Overall, the data contains both *sporadically missing values*, (variables with missing values for some patients within a dataset) and *systematically missing values* (variables that are completely missing for a whole dataset, e.g., where the datasets did not record a particular variable). For *sporadically missing* values, we will assume they are missing at random. We will assume *systematically missing values* are missing completely at random[33].

### Statistical Analysis Methods

All our analysis choices have required us to respect that we will not have direct access to the individual participant data (IPD) from all datasets; we will only be able to access some datasets through remote analysis whereby our analytical scripts are sent to the data custodians, who run them and return to us the analytical results. Specifically, we have direct (IPD-level) access to 18 of the 23 datasets, which we will “stack” into a single database (hereafter named ‘datastack’), while the remaining 5 datasets will be analysed remotely for data governance reasons (‘remote data’). To account for this approach to data access, we have developed a bespoke two-phase design, following best-practice recommendations.[34,35] We provide an overview of the statistical processes in this section. A graphical representation of our modelling approach is given in **Figure 2**.

**Figure 2:**
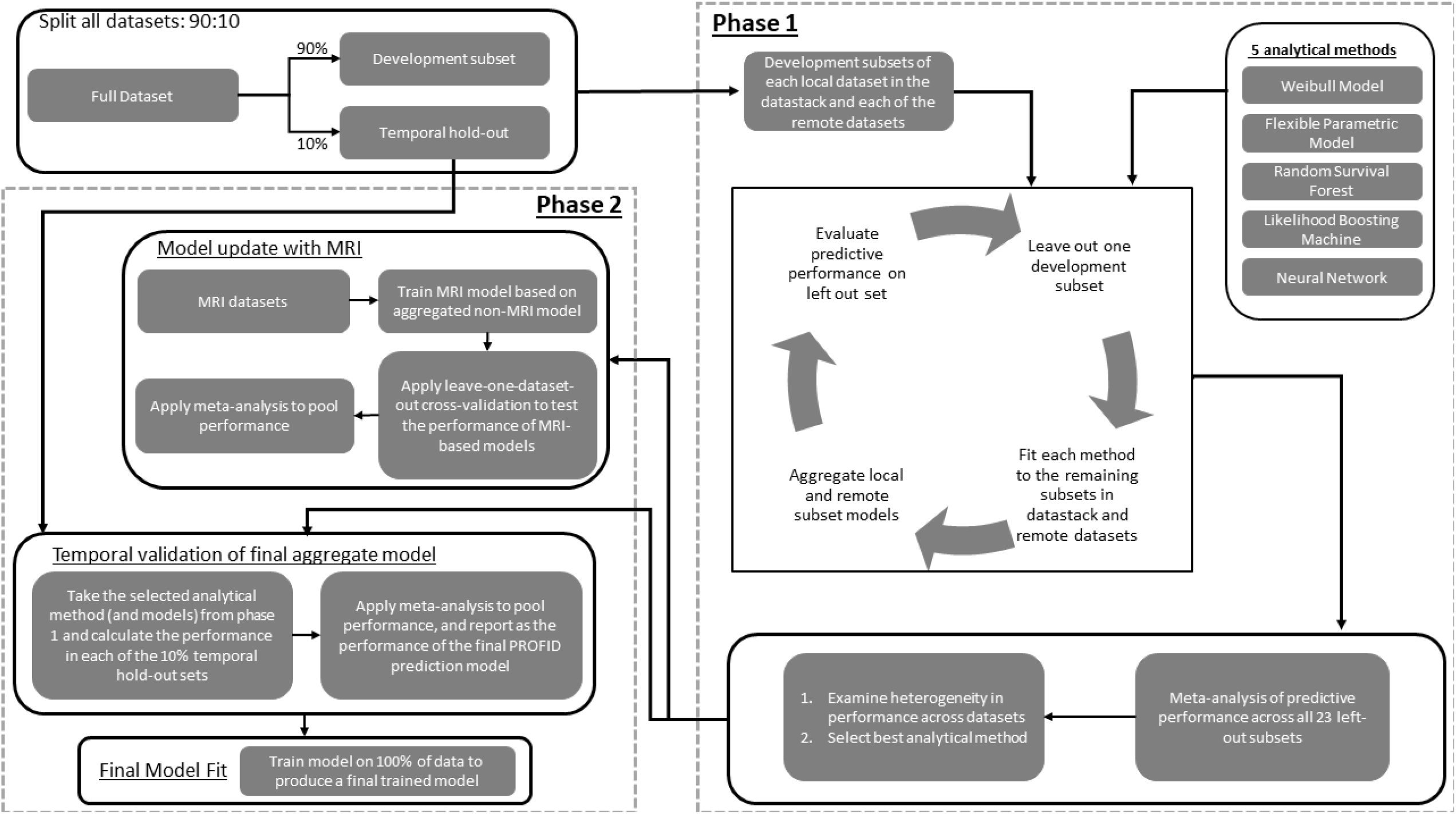
Graphical representation of our modelling approach.

In all assessments of predictive performance that we mention below, we quantify this using discrimination, calibration and overall accuracy; all measures will be estimated within the competing risk framework.[36] We will quantify discrimination of the prediction model using the weighted Harrell’s weighted C-index.[37] We will assess the calibration using calibration plots of the observed survival curves (Kaplan-Meier plot) against those predicted by the model. Where relevant (i.e. for regression-based methods), we will also summarise calibration by estimating the calibration slope (ideal value 1) by fitting a Cox regression model (to the sub-distribution hazard of SCD) to the observed outcomes and with the linear predictor (for regression models) from the model as the only covariate. Finally, we will compute overall accuracy through the integrated Brier score. The inverse probability of censoring weights versions of both Harrell’s C-index and the integrated Brier score will be based on Kaplan-Meier estimates of the censoring distribution in the training sets.

#### Phase 0: data preparation and ratification

From the onset of our analysis, within all datasets we will temporally hold-out 10% of data using an event-stratified sampling approach based on the latest event times (i.e., where we randomly select the latest 10% of the temporally ordered event dates), which is similar to period analysis [38]. Hereafter, these are all called the ‘10% hold-out sets’, with the remaining 90% of each dataset called the ‘development sets’ (**Figure 2**). The 10% hold-out sets will be used in the second phase of modelling to temporally validate the selected analytical model from phase 1 (see below). We emphasise that within each modelling phase our approach to validating CPMs is systematic internal-external cross-validation.[39,40] As a preliminary modelling step, we will undertake a data ratification exercise, where reports will be sent to each data provider to ensure consistency in approaches to cleaning and analyzing each data source.

#### Phase 1: model development, aggregation, and systematic internal-external cross-validation

For this first phase of the main analysis, we use model aggregation methods combined with systematic inter-external cross-validation (across the 90% development subsets) [39,40] to select the “best” analytical method to take forward for the final PROFID CPM (**Figure 2**). Specifically, this process involves leaving out one of the 90% development subsets, with the remaining development subsets used to develop a CPM on (a) the datastack combined and (b) on each remote dataset in turn, using each of the following methods (within a competing risk framework): 1) Weibull modelling, 2) flexible parametric survival modelling [41,42], 3) random survival forests [43], 4) likelihood boosting machine, and 5) neural network [44] (see the supplementary methods for details on how we will fit each method and variable selection). For each of the five analytical methods, this will create either 5 or 6 CPMs in each iteration of the leave-one-out-cross-validation (5 if the left-out set is a remote dataset, and 6 if the left-out set is within the datastack, since we have the datastack plus 5 other remote datasets). Since we need a single PROFID model, these 5/6 CPMs will then be combined using the methods described by Debray et al [45] (see supplementary methods) to create five aggregated CPMs (one aggregated CPM per analytical method). Each of these five aggregate models will then be validated in the left-out 90% development subset. This process then cycles through leaving out each development subset in-turn, resulting in 23 sets of predictive performance estimates per analytical method. We will use random effects meta-analysis to combine these estimates, resulting in 5 sets of pooled (meta-anlaysed) predictive performance estimates, with associated measures of heterogeneity (I^2^) and prediction intervals (i.e. the potential model performance in a new population similar to those included in the meta□analysis). [46]

At this point, we will consider leaving out some datasets if the hetrogenity assessment indicates some data are markedly different (detrimentally) to the others. Any such decisions will also be based on clinical assessments, to avoid this being a completely data-driven exercise. If we do leave out a dataset, then the aforementioned internal-external validation processes would be repeated.

After completing this step, we will use the pooled (meta-analysed) predictive performance measures to select the analytical method for the final PROFID CPM. We will favour the analytical method that leads to the highest predictive performance in terms of calibration and discrimination. If multiple methods lead to similar performance or if different performance metrics favour different models, then we will favour the model that has the fewest number of predictor variables and is most transparent in how it calculates risk estimates (interpretability) and has the easiest clinical implementation.

#### Phase 2: model updating for CMR variables and final model systematic internal-external cross-validation

At the end of phase 1, we will have selected the analytical method that will be used for the final PROFID CPM. In phase 2, we will seek to use the temporally held-out sets (10% hold-out sets) to obtain unbiased estimates of predictive performance of said model (having used the prediction performance estimates from phase 1 to select an analytical method). Specifically, we will take the selected analytical method (and corresponding models) from phase 1 and calculate the performance in each of the 10% temporal hold-out sets separately; to estimate genuine temporal performance, we will not use the 90% development subsets further here, but rather take the models as developed from phase 1. As in phase 1, we will apply meta-analysis to pool the estimates of predictive performance of the final aggregate model [46]; this pooled set of performance estimates become our final external validation measures, and we will also calculate prediction intervals for each performance estimate (i.e. the potential model performance in a new population similar to those included in the meta-analysis [47]).

Additionally, phase 2 will also consider the addition of the CMR variables into the modelling (using six datasets that have such data recorded). Having determined the best analytical approach (phase 1), we will again employ the systematic internal-external cross-validation framework on the subset of datasets with CMR variables to perform flexible parametric extension and recalibration [48–54] thereby allowing inclusion of CMR variables into the model. Specifically, we will train a flexible parametric survival model with the following covariates: the logit-transformed 1-year probability output of the selected phase 1 model, core scar estimated with the full-width half-maximum (FWHM) method, and grayzone. Maintaining appropriate data segregation to avoid information leakage, we will impute core scar in the one dataset without FWHM (Centro Cardiologico Monzino Registry), whereas we will use quantile normalisation to obtain a unified representation of grayzone across all datasets, where several different methods were used for quantification. Iterating across all leave-one-out datasets, we will then apply meta-analysis to pool predictive performance of this extended model.

Finally, using the 10% temporal hold-out sets, we will compare the 1-year predicted risks for individuals from the CPM with CMR data and the CPM without CMR data, thus providing the likelihood that the 2.5% risk threshold (for the PROFID-Reduced trial) or 3% threshold (for the PROFID-Preserved trial) be crossed after the addition of CMR given the initial PROFID CPM prediction. This will inform the decision of subgroups of patients (combinations of risk variables) where requesting CMR data at prediction time would be informative.

As a final analytical step, we will use 100% of the datastack and of each remote dataset (again combining the models across these using model aggregation) to fit the final PROFID, using the best analytical method.

### Model Output

The output of the PROFID model will be i) risk of SCD preventable by an ICD implant, corrected for risk of death by competing causes, and ii) risk of death by competing causes, corrected for risk of SCD. For context, we will also report overall mortality risk (i.e., risk of SCD + risk of death from other causes).

## Ethics and Dissemination

All patient data analysed in this study will be de-identified and stored in a secure data environment for analysis; all individual participating databases were approved by their respective ethical boards, whenever required by national or local regulations strictly in adherence to GDPR, European laws and local data safety protocols.

We will disseminate our results through scientific publications. All statistical code for the analysis will be made available through open-source repositories (e.g., git/github) for full transparency and reproducibility. We will not be able to share the individual participant data owing to the required data sharing agreements and considerations. Finally, we will embed the final model in an appropriate open-source platform (e.g., web-calculator/ app). All dissemination will follow the TRIPOD guidelines.[22]

## Data Availability

We will not be able to share the individual participant data owing to the required data sharing agreements and considerations

