## Supplementary Material for "Development and Validation of a Multivariable Risk Prediction Model for Sudden Cardiac Death after Myocardial Infarction (PROFID Risk Model): Study Rationale, Design and Protocol"

### Supplementary Methods

#### Statistical Analysis

##### Analytical Methods of Model Fitting

In the first modelling stage, we will apply five different analytical methods to develop a risk prediction model in the datastack and the remote datasets, all under a competing risk framework: 1) Weibull modelling, 2) flexible parametric survival modelling, 3) random survival forests, 4) likelihood boosting machines, and 5) neural networks. Each of these analytical methods require different considerations in terms of model fitting, which we describe as follows:

- Both the flexible parametric model and the Weibull model will be fit using *stpm2cr* Stata module [1]. For the flexible parametric survival models, the number of spline knots will be set to a single internal knot as recommended by [2] but we will also explore model fits with 1 to 3 internal knots, where possible; their locations will be at quantiles of the event times. For both the Weibull and flexible parametric models, continuous variables will be modelled as such, including consideration of non-linear effects using natural cubic splines, with 3 internal knots. Furthermore, interaction terms will be defined *a priori* based on clinical insights. Variable selection for the Weibull and flexible parametric models will be based on fitting a Cox proportional hazards model (competing risks framework) under a LASSO penalised likelihood; the LASSO penalisation parameter will be optimised using cross-validation using the one-SE method to derive the most parsimonious model. The Weibull and flexible parametric models will then be fitted using maximum likelihood with the predictor variables that were selected by the LASSO, with forward-backwards stepwise selection based on AIC then initiated to further refine the selected predictor variables. [3–5] This two-step process of variable selection is required since there is no software available to fit Fine & Gray Weibull/flexible parametric survival models under a LASSO likelihood (which would be the ideal situation), but our two-step process is designed to be a good compromise as it avoids selecting across large number of variables using stepwise methods. Regression coefficients will be shrunk, post-estimation, using heuristic shrinkage factors [6].

- For random forest models, [7] the following hyper-parameters will be optimised using integrated Brier score on the out-of-bag samples: number of trees, number of variables to try at each binary split and the minimum node size. The splitting rule will be the modified weighted log-rank test based on Gray's test.[8] The number of random splits will be set to two, as opposed to considering all possible splits. The models will be fit using randomForestSRC R package.[6,7,9–11] We will consider interactions between variables (defined *a priori* based on clinical insights, as above) by explicitly considering them in the design matrix.
- The (partial) likelihood boosting machine used in this analysis uses Fine and Gray's competing risks regression as base models with the underlying sub-distribution cumulative hazard functions estimated using Breslow-type method. The consecutive models are fit based on penalised partial likelihood. The boosting procedure results in some coefficients' values set to zero, which, effectively, results in predictor selection.[9,12] The optimal number of boosting steps and the penalty parameter will be selected using 5-fold cross-validation on the development data based on integrated Brier score. The models will be fit using the CoxBoost R package.[10] We will consider interactions between variables (defined *a priori* based on clinical insights, as above) by explicitly considering them in the design matrix.
- For the neural network models, we will use DeepHit.[13]. The model requires optimisation of many parameters and hyperparameters, as follows, which we will optimise using integrated Brier score through 5-fold cross-validation on the development data: (i) the number of layers for each sub-network, (ii) the number of neurons for each layer of each sub-network and (iii) the optimal number of variables for variable selection. Variable selection will be based on permutation importance in the development data.[14,15] The models will be fit using the following: python3, pandas, numpy, scikit_learn, lifelines, hyperopt and tensorflow. We will consider interactions between variables (defined *a priori* based on clinical insights, as above) by explicitly considering them in the design matrix.

One important consideration for all modelling is how we handle LVEF. In particular, the timing of the LVEF measurements that are recorded differs across the datasets. Specifically, some datasets, such as the Swedish Heart Registry, only include LVEF measured at the time of the MI (before the remodelling period), others include LVEF measured after the remodelling period (e.g., at time of ICD implantation), and one dataset (Silesian RDB) includes both. To allow us to incorporate all data on LVEF from all datasets (given its importance as a predictor), at model development stage we will include an interaction term between LVEF and an indicator of its timing (before/after remodelling).

All of the methods stated above have their own approaches to selecting predictor variables (from the candidate predictor variables in the common data model). Such variable selection processes only aim to maximise/optimise prediction performance. Nonetheless, it is also important that our final model is clinically acceptable in terms of the number of included predictors. Given we have approximately 100 candidate predictor variables, it is possible that large numbers of predictors will be selected for inclusion in the model, which would be unacceptable from a clinician end-user perspective. Therefore, we will also consider models fitted where the maximum number of predictor variables is capped at the 20 most “important” and models where the maximum number of predictor variables is capped at the 10 most “important”. For the Weibull and flexible parametric models, these 20/10 predictor variables will be those selected at the value of the LASSO penalisation parameter that results in 20/10 selected variables (which will then be refined using stepwise process described above), while for the machine learning models, this will be based on the top 20/10 variables according to variable importance.

#### Model Aggregation

Once we have fit models to the development sets of (a) the datastack, and (b) each remote dataset, this creates 5-6 CPMs (as documented in main paper) for each analytical method. Each of these will subsequently be combined/ aggregated per modelling method (i.e., we will not aggregate across modelling methods) using the development sets of each dataset (see **Figure 2** of main paper). We will use a generic approach to aggregate the models (i.e., an aggregation method that works for all 5 analytical methods we consider).

Specifically, we will use a weighted average of predictions across all datasets, for a given modelling approach. Taking $f_{m,j}(x_i, t)$ to denote the prediction for patient $i$ at time $t$ from model $m$ (indexed across the datastack and remote datasets) for approach $j$ (Weibull, Flexible parametric, etc.), then for each $j$ separately we will define our aggregate model to be:

$$f_j(x_i, t) = \frac{\sum_{m=1}^{M} \sqrt{n_m} f_{m,j}(x_i, t)}{\sum_{m=1}^{M} \sqrt{n_m}}$$

where $M$ is the total number of datasets across the datastack and remote data (will be either 5 or 6, as documented in main paper), and $n_m$ is the sample size of dataset $m$. That is, the aggregate models will be a weighted average of the predictions from all models, where the weights will be based on square-root of the sample size of each dataset.

### Supplementary Tables

**Supplementary Table 1**: The PROFID common data model, which shows a consistent set of variables across all data sources. In particular, the common data model dictates the units of measurement where applicable, categories for nominal and ordinal variables, and definitions of each variable.

| Classification | Variable name | Categories/Units | Description |
|---|---|---|---|
| Demographics | Age | years | Age |
| Demographics | Sex | Male/Female | Sex |
| Demographics | Ethnicity | Black/Other | Ethnicity |
| Clinical characteristics | BMI | $kg/m^2$ | Body mass index |
| Clinical characteristics | SBP | mmHg | Systolic blood pressure |
| Clinical characteristics | DBP | mmHg | Diastolic blood pressure |
| Clinical characteristics | NYHA | I/II/III/V | New York Heart Association classification |
| Clinical characteristics | MI_type | STEMI/NSTEMI | Myocardial infarction (MI) type at index MI |
| Clinical characteristics | MI_location_anterior | No/Yes | Location of index MI - anterior |
| Clinical characteristics | MI_location_inferior | No/Yes | Location of index MI - inferior |
| Clinical characteristics | MI_location_lateral | No/Yes | Location of index MI - lateral |
| Clinical characteristics | MI_location_posterior | No/Yes | Location of index MI - posterior |
| Clinical characteristics | PCI | No/Yes | PCI prior to or at the time of follow-up start |

| | | | |
|---|---|---|---|
| Clinical characteristics | CABG | No/Yes | CABG prior to or at the time of follow-up start |
| Clinical characteristics | PCI_acute | No/Yes | PCI during index MI |
| Clinical characteristics | CABG_acute | No/Yes | CABG during index MI |
| Clinical characteristics | Thrombolysis_acute | No/Yes | Thrombolysis during index MI |
| Clinical characteristics | Revascularisation_acute | No/Yes | Revascularisation during index MI (PCI/CABG/thrombolysis) |
| Clinical characteristics | Diseased_arteries_num | N/A | Number of diseased coronary arteries |
| Clinical characteristics | LVH | No/Yes | Left ventricular hypertrophy |
| ECHO measurements | LVEF | % | Left ventricular ejection fraction (echocardiography) |
| ECHO measurements | LVDD | mm | Left ventricular diastolic diameter |
| Circulating biomarkers | BUN | mmol/L | Blood urea nitrogen |
| Circulating biomarkers | Cholesterol | mg/dL | Total cholesterol |
| Circulating biomarkers | CRP | mg/L | C-reactive protein |
| Circulating biomarkers | eGFR | $mL/min/1.73m^2$ | Estimated glomerular filtration rate (CKD-EPI) |
| Circulating biomarkers | Haemoglobin | g/dL | Haemoglobin |
| Circulating biomarkers | HbA1c | % | Haemoglobin A1c |
| Circulating biomarkers | HDL | mg/dL | High-density lipoprotein cholesterol |
| Circulating biomarkers | IL6 | pg/mL | Serum interleukin-6 |
| Circulating biomarkers | LDL | mg/dL | Low-density lipoprotein cholesterol |

| | | | |
|---|---|---|---|
| Circulating biomarkers | NTProBNP | pmol/L | N-terminal pro-BNP |
| Circulating biomarkers | Potassium | mmol/L | Potassium |
| Circulating biomarkers | Sodium | mmol/L | Sodium |
| Circulating biomarkers | Triglycerides | mg/dL | Triglycerides |
| Circulating biomarkers | Troponin_T | ng/l | Troponin T |
| Circulating biomarkers | TSH | mU/L | Thyroid-stimulating hormone |
| ECG | HR | bpm | Resting heart rate (12-lead ECG) |
| ECG | PR | ms | PR interval (12-lead ECG) |
| ECG | QRS | ms | QRS duration (12-lead ECG) |
| ECG | QTc | ms | Corrected QT interval (Bazett; 12-lead ECG) |
| ECG | AV_block | No/Yes | Atrioventricular block |
| ECG | AV_block_II_or_III | No/Yes | AV block second or third degree |
| ECG | LBBB | No/Yes | Left bundle branch block |
| ECG | RBBB | No/Yes | Right bundle branch block |
| Holter monitor | HR_average | bpm | Average heart rate |
| Holter monitor | SDNN | ms | Standard deviation of NN intervals |
| Medical history | MI_history | No/Yes | History of prior MI before index MI |
| Medical history | Time_1st_MI | months | Time since first MI |

| | | | |
|---|---|---|---|
| Medical history | Time_index_MI | months | Time since index MI |
| Medical history | HF | No/Yes | Heart failure |
| Medical history | Stroke_TIA | No/Yes | Stroke or transient ischaemic attack |
| Medical history | NSVT | No/Yes | History of non-sustained ventricular tachycardia |
| Medical history | AF | No/Yes | Atrial fibrillation |
| Medical history | Atrial_flutter | No/Yes | Atrial flutter |
| Medical history | AF_atrial_flutter | No/Yes | Atrial fibrillation or atrial flutter |
| Medical history | Anaemia | No/Yes | Anaemia |
| Medical history | Cancer | No/Yes | Cancer |
| Medical history | Pulmonary_disease | No/Yes | Pulmonary disease including COPD |
| Medical history | COPD | No/Yes | Chronic obstructive pulmonary disease |
| Medical history | Pulmonary_embolism | No/Yes | Pulmonary embolism |
| Medical history | Dementia | No/Yes | Dementia |
| Medical history | Diabetes | No/Yes | Diabetes |
| Medical history | Hyperlipidaemia | No/Yes | Hyperlipidaemia |
| Medical history | Hypertension | No/Yes | Hypertension |
| Medical history | Liver_disease | No/Yes | Liver disease |
| Medical history | Renal_disease | No/Yes | Renal disease |

| | | | |
|---|---|---|---|
| Medical history | VD | No/Yes | Evidence of any valve disease |
| Medical history | Valve_RR | No/Yes | Evidence of repair or replacement of any valve |
| Family history | FH_CAD | No/Yes | Family history of CAD |
| Family history | FH_SCD | No/Yes | Family history of SCD |
| Life style | Alcohol | No/Yes | Alcohol consumption |
| Life style | Smoking | No/Yes | Smoking |
| Cardiac MRI | MRI_LVEF | % | LVEF based on cardiac MRI |
| Cardiac MRI | Infarct_size | g | Infarct size (core scar) |
| Cardiac MRI | Greyzone_size | g | Greyzone size |
| Cardiac MRI | Total_scar | g | Total scar: infarct size + greyzone |
| Cardiac MRI | LV_mass | g | Left ventricular mass |
| Cardiac MRI | LVEDV | mL | Left ventricular end diastolic volume |
| Cardiac MRI | LVESV | mL | Left ventricular end systolic volume |
| Cardiac MRI | MRI_time_baseline | months | Time interval between cardiac MRI and baseline |
| Year of baseline | Time_zero_Y | N/A | Calendar year of follow-up start |
| Medication | ACE_inhibitor_ARB | No/Yes | ACE inhibitor<br><br>ARB: Angiotensin receptor blocker |

| | | | |
|---|---|---|---|
| Medication | ACE_inhibitor | No/Yes | ACE inhibitor |
| Medication | ARB | No/Yes | Angiotensin receptor blocker |
| Medication | Aldosterone_antagonist | No/Yes | Aldosterone antagonist also known as mineralocorticoid antagonist |
| Medication | Anti_anginal | No/Yes | Anti-anginal |
| Medication | Anti_arrhythmic | No/Yes | Anti-arrhythmic |
| Medication | Anti_arrhythmic_III | No/Yes | Anti arrhythmic class III |
| Medication | Anti_arrhythmic_1C | No/Yes | Anti-arrhythmic class 1C |
| Medication | Anti_coagulant | No/Yes | Oral anti-coagulant |
| Medication | Anti_coagulant_nonVKA | No/Yes | Oral anti-coagulant non-VKA |
| Medication | Anti_coagulant_VKA | No/Yes | Oral anti-coagulant VKA |
| Medication | Anti_diabetic | No/Yes | Anti-diabetic |
| Medication | Anti_diabetic_insulin | No/Yes | Anti-diabetic: Insulin |

| | | | |
|---|---|---|---|
| Medication | Anti_diabetic_oral | No/Yes | Oral anti-diabetic |
| Medication | Anti_platelet | No/Yes | Anti-platelet |
| Medication | Beta_blockers | No/Yes | Beta-blocker |
| Medication | Calcium_antagonists | No/Yes | Calcium antagonists |
| Medication | Digitalis_glycosides | No/Yes | Digitalis glycosides |
| Medication | Diuretics | No/Yes | Diuretic |
| Medication | Lipid_lowering_other | No/Yes | Lipid lowering medication others than statins |
| Medication | Lipid_lowering_statins | No/Yes | Statins |
